# A Mendelian randomization study identifies proteins involved in neurodegenerative diseases

**DOI:** 10.1101/2024.02.24.24303314

**Authors:** Lazaros Belbasis, Sam Morris, Cornelia van Duijn, Derrick Bennett, Robin Walters

## Abstract

Proteins are involved in multiple biological functions. High-throughput technologies have allowed the measurement of thousands of proteins in population biobanks. In this study, we aimed to identify proteins related to Alzheimer’s disease (AD), Parkinson’s disease (PD), Multiple Sclerosis (MS) and Amyotrophic Lateral Sclerosis (ALS) using large-scale genetic and proteomic data.

We performed a two-sample *cis* Mendelian randomization (MR) study by selecting instrumental variables for the abundance of over 2,700 proteins measured by either Olink or SomaScan platforms in plasma from UK Biobank and the deCODE Health Study. We also used the latest publicly-available GWAS for the diseases of interest. The potentially causal effect of proteins on neurodegenerative diseases was estimated based on the Wald ratio.

We tested 10,244 protein–disease associations, identifying 122 associations which were statistically significant (5% false discovery rate). Out of 57 associations (58%) tested using an instrumental variable from both Olink and SomaScan platforms, 33 (58%) were statistically significant in both platforms. Evidence of co-localisation between plasma protein abundance and disease risk (posterior probability >0.80) was identified for 46 protein-disease pairs. Twenty-three out of 46 protein–disease associations correspond to genetic loci not previously reported by genome-wide association studies.

The newly-associated proteins for AD are involved in complement (*C1S*, *C1R*), microglia (*SIRPA*, *PRSS8*) and lysosomal functions (*CLN5*). A protein newly-associated with PD (*CTF1*) is involved in the interleukin-6 pathway, two proteins for ALS (*TPP1*, *TNFSF13*) are involved in lysosomal and astrocyte function, respectively, and proteins associated with MS are involved in blood–brain barrier function (*TYMP*, *VEGFB*), the oligodendrocyte function (*PARP1*), the structure of the node of Ranvier and function of dorsal root ganglion (*NCS1*, *FLRT3*, *CDH15*), and the response to viral infections including Epstein-Barr virus (*PVR*, *WARS1*). Our study demonstrates how harnessing large-scale genomic and proteomic data can yield novel insights into the role of plasma proteome in the pathogenesis of neurodegenerative diseases.

## Introduction

Neurological diseases are the leading cause of disability and the second leading cause of death worldwide.^1^ Neurodegenerative diseases constitute a distinct group of neurological diseases, which are characterised by progressive neuronal loss and formation of distinct pathological changes in the brain.^2^ During the last three decades, there has been a substantial increase in the number of people living with neurodegenerative diseases such as Alzheimer’s Disease (AD), Parkinson’s Disease (PD), Amyotrophic Lateral Sclerosis (ALS), and Multiple Sclerosis (MS).^3–6^ Genome-wide association studies (GWAS) have identified molecular pathways leading to neurodegenerative diseases and have increased our knowledge on causal pathways involved in these diseases.^7–10^

Proteins play a key role in a range of biological processes, so that their dysregulation can lead to the development of diseases and even minor modulation of their levels or function can modify disease risk. They represent a major source of biomarkers for the diagnosis or prediction of disease, and may also be crucial to improving our understanding of the pathogenesis of diseases.^11^ About 75% of FDA-approved medications were targeted at human proteins.^12,13^ Therefore, by combining large-scale genomic and proteomic profiling, there is a potential to identify disease-causing pathways, uncover new drug targets, highlight novel therapeutic indications, and identify clinically relevant biomarkers.^14,15^

Recent technological advances have allowed the measurement of thousands of proteins in large population-based studies. To date, two different high-throughput techniques to measure the abundance of multiple proteins have been used in large population samples: an antibody-based proximity-extension assay (*Olink* platform) and an aptamer affinity-based assay (*SomaScan* platform).^15^ GWAS of plasma protein abundance have identified protein quantitative trait loci (pQTLs), which can be used to examine the potentially causal effect of proteins on human diseases and traits using the Mendelian randomization (MR) framework.^11^ MR is an instrumental variable (IV) approach, which can be used to accelerate the discovery of biomarkers and the drug development pipeline.^16^ MR studies have examined the potential role of proteins in the development of neurological diseases, by mainly following a transcriptome-wide MR approach, which uses expression quantitative trait loci (eQTLs) as IVs; ^17–22^ however, the value of such analyses is limited by the fact that eQTLs frequently do not accurately reflect protein abundance.^23^

In the present study, we harnessed summary-level genetic data from two large proteo-genomic studies and from the largest GWAS for major neurodegenerative diseases, to identify proteins whose abundance in plasma is associated with these diseases (**Figure 1**). We followed a two-sample *cis* MR approach, and we minimised the risk of confounding by linkage disequilibrium (LD) by performing a co-localisation analysis. We complemented our analysis by exploring the potential effects of these proteins on multiple brain imaging phenotypes.

**Figure 1.**
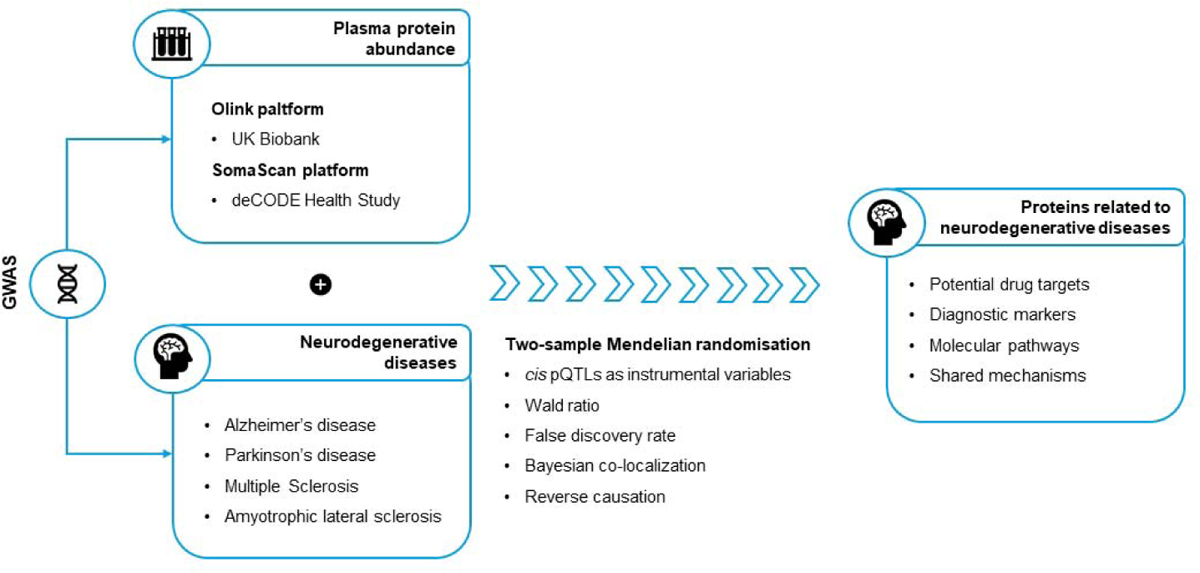
Schematic representation of the study. *cis* protein quantitative trait loci (pQTLs) from two large proteogenomic studies were used as instrumental variables for protein abundance in plasma. The association of plasma proteins with neurodegenerative diseases was assessed by estimating the Wald ratio in a two-sample Mendelian randomisation framework. The analyses were complemented by Bayesian co-localisation.

## Materials and methods

### Data sources

#### GWAS of human plasma proteome

We used summary-level data from the two largest proteogenomics studies conducted in populations of European ancestry using either *Olink* or *SomaScan* platforms, which were identified through a publicly available catalogue of proteogenomics studies (last update on 29^th^ January 2024).^15^ The association between circulating protein levels and genetic variants was assessed in:

- 35,571 participants in the UK Biobank using the Olink Explore 1536 platform, measuring 2,941 protein analytes, capturing 2,923 unique proteins,^24^ and
- 35,559 participants in the Icelandic Cancer Project and deCODE Health Study using the SomaScan version 4 platform, measuring 4,907 aptamers, capturing 4,719 proteins.^25^

We do not assume equivalence of the two platforms, but we included pQTLs identified through both platforms to increase the completeness of our analysis.

#### GWAS of neurological diseases

We searched GWAS Catalog for published GWAS on neurological diseases (**Supplementary Table 1**).^26^ We selected the largest publicly available GWAS in population of European ancestry for Alzheimer’s Disease (111,326 cases, 75 genome-wide significant variants),^8^ Parkinson’s Disease (33,674 cases, 90 genome-wide significant variants),^7^ Multiple Sclerosis (47,429 cases, 200 genome-wide significant variants),^10^ and Amyotrophic Lateral Sclerosis (27,205 cases, 15 genome-wide significant variants).^9^

### Selection of instrumental variables for protein abundance

pQTLs are genetic variants with an effect on protein expression, and they could be either *cis* or *trans* based on their proximity to the gene encoding the protein of interest.^27^ *trans* pQTLs map to genes that do not directly code for the targeted proteins or that correspond to intergenic regions, and it is difficult to distinguish between detected effects due to vertical and horizontal pleiotropy.^11,28^ For this reason, we restricted our Mendelian randomisation (MR) analysis to *cis* pQTLs. Only autosomal genetic variants were included in the analyses, because summary statistics for chromosome X are not available in some of the GWAS for neurological diseases.

We retrieved statistically significant *cis* pQTLs from each of the proteogenomics studies, applying the same level of statistical significance as used in those studies (*P* < 3.40 × 10^-11^ for the study using the Olink platform, and *P* < 1.80 × 10^-9^ for the study using the SomaScan platform). For Olink platform, the *cis* region was defined as a distance of 1 Mb upstream or downstream from the gene encoding the protein of interest.^24^ For SomaScan platform, the *cis* region was defined as a distance of 1 Mb upstream or downstream from the transcription start site of the gene encoding the protein of interest.^25^

We used the following criteria to filter the list of statistically significant *cis* pQTLs in each study:

1. Due to the complex linkage disequilibrium (LD) structure of SNPs within the human major histocompatibility complex (MHC) region, SNPs and proteins encoded by genes within the MHC region (chr6: from 26 Mb to 34 Mb) were excluded.
2. To reduce the risk for weak instrument bias, we calculated the F-statistic for each SNP, and we excluded genetic instruments with an *F*-statistic <10.^29^ Details on the estimation of *F*-statistic are presented in the Supplementary Material.
3. We obtained the genetic variants that were also tested in the GWAS for neurological diseases.
4. For each protein we selected only the cis pQTL with the lowest *P*-value, which we refer as the “lead variant”.

As our main analysis, we aimed to select non-overlapping instruments from each proteo-genomics study. We mapped all the proteins to UniProt IDs. When multiple assays in the same platform targeted the same protein (as defined by UniProt ID), we included only the instrument with the lowest *P*-value. For proteins with a *cis* pQTL available in both platforms, we considered only the *cis* pQTL from the UK Biobank Olink study.

### Data harmonization

All GWAS summary statistics were lifted over to genomic build 38.^30^ We followed the recommended harmonisation framework for two-sample MR analyses.^31,32^ Ambiguous palindromic single nucleotide polymorphisms with an allele frequency between 0.42 and 0.58 were excluded to avoid potential allele mismatch across different GWAS.^31^ Data harmonization was implemented using the *TwoSampleMR* package through in-built functions.^33,34^

### Statistical analysis

#### Association of protein abundance with neurological diseases

The Wald ratio, which is defined as the ratio of the gene-outcome effect divided by the gene-exposure effect, was calculated for all the protein-disease associations.^35^ To identify the statistically significant associations, a multiplicity correction was applied using the Benjamini-Hochberg method.^36^ Evidence of a statistically significant protein-disease association was based on 5% false discovery rate (FDR). MR analyses were performed using the *TwoSampleMR* package.^33,34^ We prioritised proteins with a statistically significant association with a neurological disease for further analyses to: a) assess reverse causality; and b) perform Bayesian co-localisation.

#### Assessment of reverse causation

Reverse causation could be a potential explanation for positive findings in an MR analysis. We explored the potential for reverse causality using a bi-directional MR approach. We performed LD clumping to obtain approximately independent genetic variants to model the genetic liability to AD, PD, ALS, and MS. Clumping was performed using the reference panel from 1000Genomes for population of European ancestry setting a statistical significance threshold of *P* < 5 × 10^-8^, a genetic window of 1Mb and an LD *r*^2^ < 0.1%. We used the *ld_clump* function from the *ieugwas* package and PLINK version 1.90.^37^ We derived four genetic instruments consisting of genome-wide significant genetic variants as reported in the relevant publications.^7–10^ We examined whether the genetically predicted liability to each one of the neurological diseases of interest was related to the proteins associated with each one of the diseases. We estimated the Wald ratio for each one of the genetic instruments and we combined them using a fixed-effect inverse-variance weighted model.^38^ Evidence of statistically significant findings were based on 5% FDR.

#### Bayesian co-localisation

Evidence of co-localisation supports the validity of the IVs and strengthen the MR findings.^39^ To assess potential confounding by LD, we examined whether the prioritised proteins share the genetic variant with the outcomes of interest by conducting a co-localisation analysis assuming a single causal variant in each genetic locus. We used the *coloc* package for the co-localisation analysis. Variants within ±1 Mb window around the pQTLs with the smallest *P*-value were included. We used a posterior probability higher than 80% as strong evidence of co-localisation, and a posterior probability higher than 60% as moderate evidence of co-localisation. However, we acknowledge that lack of co-localisation does not invalidate the MR findings, as co-localisation methods have a high false negative rate.^12,40^ The GWAS by Bellenguez et al^8^ for AD do not provide the majority of the genetic variants in the *APOE* gene locus. For this reason, we repeated the co-localization analysis using the GWAS by Kunkle et al^41^ for the genetic loci located nearby *APOE* (i.e., *APOE*, *APOC1*, and *NECTIN2*).

#### Association of mRNA abundance with neurological diseases

GWAS of gene expression reported *cis* eQTLs which are genetic variants affecting the messenger RNA (mRNA) abundance.^42^ The *eQTLGen* consortium examined eQTLs from blood-derived expression of 19,250 autosomal genes and reported at least one *cis* eQTL for 16,987 genes using a sample of 31,684 individuals.^43^ The *MetaBrain* consortium provides *cis* eQTLs in 5 tissues (cortex [2,683 individuals], cerebellum [492 individuals], basal ganglia [208 individuals], hippocampus [168 individuals], and spinal cord [108 individuals]).^44^ For each one of the statistically significant proteins in the previous step, the relevant lead *cis* eQTL was selected as a genetic IV. We used the same statistical significance threshold as the GWAS on plasma and brain eQTLs to identify appropriate genetic instruments. We estimated the Wald ratio as the ratio of the genetic effect on disease risk divided by the genetic effect on mRNA abundance. A multiplicity correction was applied using the Benjamini-Hochberg method separately in plasma and brain tissues,^36^ and statistically significant associations were assessed at 5% FDR.

#### Association of protein abundance with brain imaging phenotypes

The potentially causal effect of the prioritised proteins on brain imaging traits was examined using summary-level GWAS data for nine brain volumes, mean cortical thickness and surface, and white matter hyper-intensities.^45–47^ The available brain volumes were intracranial volume,^48^ hippocampal volume,^49^ and other subcortical structures volume^50^ (nucleus accumbens, amygdala, brainstem, caudate nucleus, globus pallidus, putamen, and thalamus).

A multiplicity correction was applied using the Benjamini-Hochberg method,^36^ and statistically significant findings were assessed at 5% FDR. Additionally, for the statistically significant associations, we performed Bayesian co-localisation analysis assuming a single causal variant and using the same specifications as described before.

## Results

### Instrumental variables for plasma protein abundance

An overview of the study design is presented in **Figure 2**. Descriptive characteristics of the data sources used in this study are shown in **Supplementary Table 1**. To identify genetic instruments of protein abundance, we used the two largest GWAS of plasma protein abundance in European populations.^25,51^ The first was conducted in the UK Biobank using the Olink Explore 1536 platform and examined 2,941 unique proteins. The second one was conducted in the deCODE Health Study using the SomaScan version 4 platform and examined 4,907 unique proteins. A *cis* pQTL was available for 2,733 unique proteins in total from either UK Biobank or deCODE Health Study. Where different instruments were available for both platforms, we used the instrument from the UK Biobank Olink study. All the IVs had an *F*-statistic > 10, minimising the influence of weak instrument bias on the MR estimates.^52^

**Figure 2.**
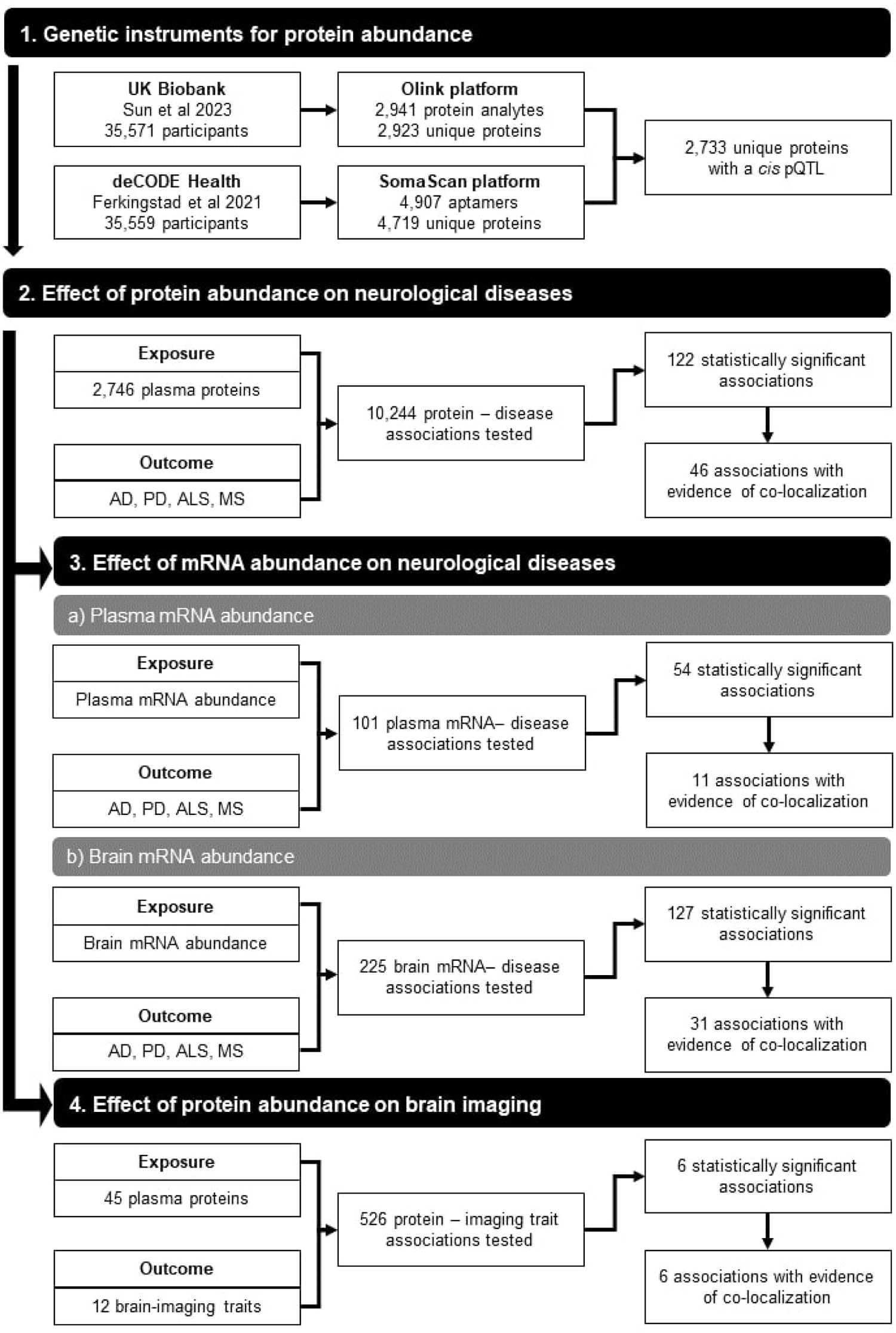
Overview of the study design and main results. The study consists of four main steps. In the first step, we identified the lead cis pQTL for 2,746 non-overlapping proteins from GWAS using the Olink and SomaScan platform in European populations. In the second step, we used these pQTLs as instrumental variables to perform a two-sample Mendelian randomisation (MR) analysis for Alzheimer’s disease (AD), Parkinson’s Disease (PD), Multiple Sclerosis (MS) and Amyotrophic Lateral Sclerosis (ALS). In the third step, we repeated the MR analysis by using plasma and brain eQTLs as instrumental variables. In the fourth step, we examined whether proteins are associated with 12 brain-imaging traits.

### Association of plasma protein abundance with neurological diseases

To systematically evaluate the evidence for a causal effect of 2,733 proteins on four neurological diseases (AD, PD, ALS, and MS), we undertook a proteome-wide two-sample MR. Overall 10,244 protein-disease associations were tested (67% using *cis* pQTLs derived using Olink platform measurements and 33% using *cis* pQTLs from the SomaScan platform; **Table 1** and **Supplementary Table S2**). We observed 988 (9.6%) nominally significant protein-disease associations at *P* < 0.05, constituting a substantial excess compared to the number expected under the null. Of these, 122 protein-disease associations (1.2%) remained statistically significant at 5% FDR, corresponding to *P* < 5.8 × 10^-4^ (**Table 2**, **Figure 3**). Even after exclusion of these associations, the remaining associations displayed substantial inflation compared to the null (**Table 1**).

**Figure 3.**
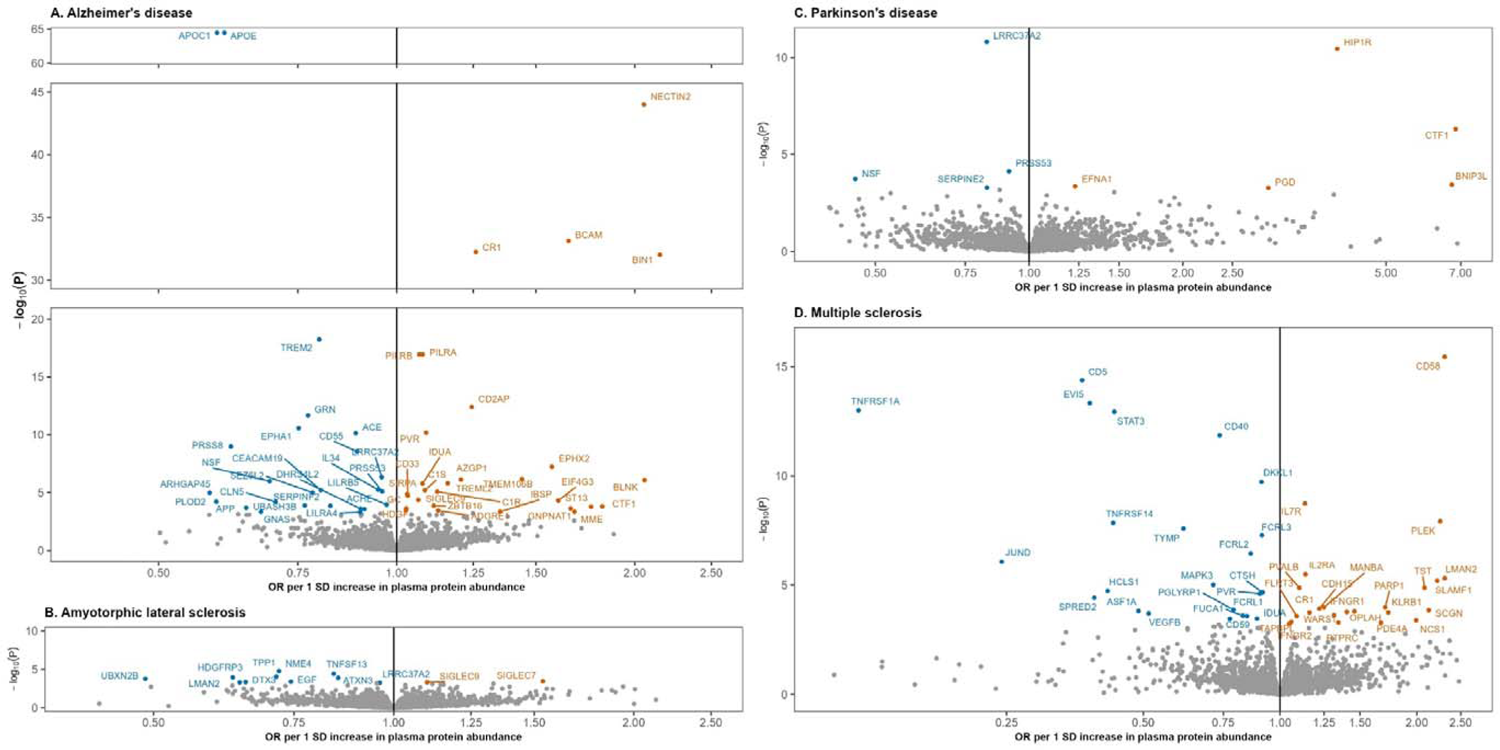
Association of proteins with neurological diseases using two-sample MR. The lead cis protein quantitative trait locus was used as an instrumental variable (IV) for 2,746 proteins. We tested 10,244 protein–disease associations, and 122 of these are statistically significant (5% FDR) and are annotated in this figure. The odds ratio corresponds to the Wald ratio, which is calculated by dividing the genetic effect of the IV on the disease by the genetic effect of the IV on the plasma protein abundance. For convenience, the statistically significant association between PTPRJ (OR = 48, P = 3.05 × 10^-4^) and Multiple Sclerosis is not shown.

**Table 1.** Summary of the proteome-wide two-sample Mendelian randomization analysis on neurological diseases.

| Disease | N associations<br>tested | N statistically significant associations |  | Inflation<br>factor<sup>2</sup> | Co-localisation<sup>3</sup> |
| --- | --- | --- | --- | --- | --- |
|  |  | P < 0.05 | 5% FDR<sup>1</sup> |  |  |
| Alzheimer's disease | 2,733 | 314 (11%) | 54 (2.0%) | 1.39 | 19 |
| Parkinson's disease | 2,202 | 167 (8%) | 9 (0.4%) | 1.11 | 2 |
| Amyotrophic lateral sclerosis | 2,663 | 213 (8%) | 12 (0.5%) | 1.28 | 2 |
| Multiple sclerosis | 2,646 | 294 (11%) | 47 (1.8%) | 1.42 | 23 |
| Total | 10,244 | 988 (10%) | 122 (1.2%) | 1.30 | 46 |

<sup>1</sup>False Discovery Rate based on Benjamini-Hochberg correction, which corresponds to a P < 5.80E-04. <sup>2</sup>Inflation factor estimated after removing the associations at 5% FDR. <sup>3</sup>Posterior probability for H4 greater than 80%. Abbreviations: FDR, false discovery rate.

**Table 2.** Proteins associated with risk of neurodegenerative diseases identified using two-sample Mendelian randomization (5% FDR) and supported by co-localisation (posterior probability >0.80).

| Assay ID<sup>1</sup> | Protein name | Genetic position<sup>2</sup> | rsID | EA/OA<sup>3</sup> | EAF | OR (95% CI)<sup>4</sup> | P | Novel<sup>5</sup> |
| --- | --- | --- | --- | --- | --- | --- | --- | --- |
| **Alzheimer's disease** |  |  |  |  |  |  |  |  |
| OID30727 | APOE | 19:44928401 | rs8106813 | G/A | 0.494 | 0.61 (0.57 - 0.64) | 3.37E-65 | No |
| OID30697 | CR1 | 1:207577223 | rs679515 | T/C | 0.174 | 1.26 (1.21 - 1.31) | 5.65E-33 | No |
| OID20197 | PILRB | 7:100374211 | rs1859788 | G/A | 0.681 | 1.07 (1.05 - 1.08) | 1.16E-17 | No |
| OID21129 | PILRA | 7:100374211 | rs1859788 | G/A | 0.681 | 1.08 (1.06 - 1.10) | 1.16E-17 | No |
| OID20177 | CD2AP | 6:47627419 | rs1385742 | A/T | 0.355 | 1.24 (1.17 - 1.32) | 3.99E-13 | No |
| OID21159 | GRN | 17:44352876 | rs5848 | C/T | 0.726 | 0.77 (0.72 - 0.83) | 2.19E-12 | No |
| OID20763 | PRSS8 | 16:31111250 | rs889555 | T/C | 0.282 | 0.62 (0.53 - 0.72) | 1.07E-09 | Yes |
| 8687_26 | TMEM106B | 7:12229791 | rs3173615 | C/G | 0.598 | 1.44 (1.25 - 1.66) | 6.93E-07 | No |
| OID30541 | BLNK | 10:96246079 | rs55769428 | A/C | 0.962 | 2.06 (1.54 - 2.74) | 8.55E-07 | No |
| OID20809 | IL34 | 16:70660097 | rs4985556 | C/A | 0.878 | 0.95 (0.93 - 0.97) | 5.65E-06 | No |
| OID30731 | C1S | 12:7063032 | rs12146727 | G/A | 0.865 | 1.09 (1.05 - 1.12) | 6.41E-06 | Yes |
| OID30753 | C1R | 12:7068900 | rs10849546 | G/A | 0.865 | 1.13 (1.07 - 1.19) | 8.55E-06 | Yes |
| OID20304 | SIRPA | 20:1915642 | rs6136377 | A/G | 0.618 | 1.03 (1.02 - 1.04) | 1.37E-05 | Yes |
| OID21472 | CD33 | 19:51225385 | rs2455069 | G/A | 0.427 | 1.03 (1.02 - 1.05) | 1.83E-05 | No |
| OID21390 | SIGLEC9 | 19:51125272 | rs2075803 | A/G | 0.447 | 1.06 (1.03 - 1.10) | 4.30E-05 | Yes |
| 6923_1 | PLOD2 | 3:146106560 | rs148118826 | G/A | 0.997 | 0.59 (0.46 - 0.76) | 6.18E-05 | Yes |
| 8874_53 | CLN5 | 13:77001261 | rs700363 | G/A | 0.910 | 0.70 (0.59 - 0.84) | 6.51E-05 | Yes |
| OID21205 | ZBTB16 | 11:114082900 | rs73000929 | A/G | 0.037 | 1.11 (1.05 - 1.18) | 1.37E-04 | Yes |
| OID21307 | MME | 3:155067802 | rs79837905 | A/G | 0.920 | 1.68 (1.26 - 2.24) | 4.54E-04 | No |
| **Parkinson's disease** |  |  |  |  |  |  |  |  |
| OID31141 | HIP1R | 12:122842051 | rs10847864 | T/G | 0.359 | 4.01 (2.66 - 6.05) | 3.64E-11 | No |
| OID20061 | CTF1 | 16:30966478 | rs11150601 | A/G | 0.628 | 6.84 (3.23 - 14.46) | 4.92E-07 | Yes |
| **Amyotrophic lateral sclerosis** |  |  |  |  |  |  |  |  |
| OID20750 | TPP1 | 11:6626391 | rs11827437 | C/T | 0.370 | 0.72 (0.62 - 0.83) | 1.61E-05 | Yes |
| OID20733 | TNFSF13 | 17:7559652 | rs3803800 | A/G | 0.211 | 0.84 (0.77 - 0.91) | 3.59E-05 | Yes |
| **Multiple sclerosis** |  |  |  |  |  |  |  |  |
| OID20716 | CD58 | 1:116547871 | rs10801908 | C/T | 0.880 | 2.30 (1.88 - 2.81) | 3.54E-16 | No |
| OID21449 | CD5 | 11:61026250 | rs4939491 | G/A | 0.609 | 0.37 (0.29 - 0.47) | 4.28E-15 | No |
| OID30519 | EVI5 | 1:92607671 | rs11808092 | C/A | 0.745 | 0.38 (0.30 - 0.49) | 4.70E-14 | No |
| OID21155 | TNFRSF1A | 12:6330843 | rs1800693 | T/C | 0.597 | 0.12 (0.07 - 0.21) | 1.02E-13 | No |
| 10346_5 | STAT3 | 17:42378745 | rs4796791 | C/T | 0.589 | 0.43 (0.35 - 0.54) | 1.17E-13 | No |
| OID20724 | CD40 | 20:46119308 | rs4810485 | G/T | 0.752 | 0.74 (0.68 - 0.80) | 1.41E-12 | No |
| OID21313 | DKKL1 | 19:49365794 | rs2303759 | T/G | 0.748 | 0.91 (0.88 - 0.94) | 1.89E-10 | No |
| OID21136 | IL7R | 5:35874473 | rs6897932 | C/T | 0.729 | 1.13 (1.09 - 1.18) | 1.84E-09 | No |
| OID20234 | TYMP | 22:50525724 | rs131805 | C/T | 0.783 | 0.61 (0.52 - 0.73) | 2.61E-08 | No |
| 9468_8 | LMAN2 | 5:177353544 | rs4131289 | A/G | 0.379 | 2.30 (1.61 - 3.30) | 4.85E-06 | Yes |
| OID20496 | SLAMF1 | 1:160668460 | rs7535367 | G/T | 0.861 | 2.22 (1.57 - 3.13) | 6.24E-06 | No |
| OID20868 | TST | 22:36862461 | rs4821544 | T/C | 0.697 | 2.08 (1.50 - 2.89) | 1.31E-05 | Yes |
| OID21420 | PVALB | 22:36862461 | rs4821544 | T/C | 0.697 | 1.10 (1.05 - 1.15) | 1.31E-05 | Yes |
| OID21011 | PVR | 19:44643102 | rs2301274 | T/C | 0.759 | 0.90 (0.86 - 0.95) | 2.46E-05 | Yes |
| OID30423 | SPRED2 | 2:65429835 | rs7569084 | C/T | 0.414 | 0.39 (0.25 - 0.61) | 3.76E-05 | No |
| OID20500 | PARP1 | 1:226421638 | rs1433574 | A/C | 0.839 | 1.70 (1.30 - 2.23) | 1.04E-04 | Yes |
| OID21005 | CDH15 | 16:89132700 | rs11646135 | A/G | 0.142 | 1.22 (1.10 - 1.35) | 1.19E-04 | Yes |
| 18172_71 | ASF1A | 6:118811617 | rs4946366 | T/C | 0.164 | 0.49 (0.34 - 0.71) | 1.53E-04 | Yes |
| OID30697 | CR1 | 1:207577223 | rs679515 | T/C | 0.174 | 1.16 (1.07 - 1.26) | 1.81E-04 | Yes |
| OID30554 | VEGFB | 11:64275525 | rs660442 | A/G | 0.200 | 0.51 (0.36 - 0.73) | 1.98E-04 | Yes |
| OID21084 | WARS | 14:100379120 | rs12882934 | C/A | 0.745 | 1.31 (1.14 - 1.52) | 2.36E-04 | Yes |
| 13123_3 | FLRT3 | 20:14693192 | rs1932953 | T/G | 0.270 | 1.09 (1.04 - 1.14) | 2.60E-04 | Yes |
| OID21458 | NCS1 | 9:130233489 | rs1054879 | A/G | 0.508 | 1.99 (1.36 - 2.92) | 4.06E-04 | Yes |

<sup>1</sup>Assay identifier as provided by the Olink or SomaScan platform. <sup>2</sup>Chromosome and genetic position in build 38 for the genetic instrumental variable. <sup>3</sup>The alleles have been orientated to reflect an increase in plasma protein abundance. <sup>4</sup>The odds ratio correspond to risk for neurological disease per 1-SD increase in plasma protein abundance estimated using the Wald ratio method. <sup>5</sup>A genetic loci was considered novel if it was not reported by the corresponding genome-wide association study. Abbreviations: CI, confidence interval; EA, effect allele; FDR, false discover rate; OA, other allele; OR, Odds ratio

One hundred and seven proteins (91%) were associated with only one neurological disease. In our study, 54 proteins associated with AD, and nine of them (17%) showed an association with AD and an additional neurological disease (CTF1, NSF, PRSS53 and PRSS8 with PD; SIGLEC9 with ALS; CR1, IDUA, and PVR with MS; and LRRC37A2 with both PD and ALS). LMAN2 showed an association with MS and ALS. Associations between proteins and phenotypes in the MR framework may reflect causality but potential alternative explanations are reverse causality, confounding by LD or horizontal pleiotropy.^11^ We evaluate each of these explanations below.

### Reverse causation

To explore the potential for reverse causation, we performed a bi-directional MR analysis, which examines whether the genetic liability to the outcome is associated with the exposure of interest. For this reason, we performed clumping to identify independent genome-wide significant variants (*P* <5.00 × 10^-8^) from the GWAS for AD, PD, ALS and MS, for use as IVs modelling the genetic liability to these diseases. Using a fixed-effect inverse-variance weighted model, we found that genetically predicted risk for AD was associated with plasma protein abundance of *APOE* and *CEACAM19* and genetically predicted risk for MS was associated with plasma protein abundance of *CD5*, *KLRB1*, *SCGN* and *MANBA* (5% FDR; **Supplementary Table S3**).

### Cross-platform comparison of cis pQTL associations

We hypothesise that a consistent causal effect in both Olink and SomaScan platforms strengthens confidence in the instruments and in the robustness of the protein–disease associations, by showing that the inferred effect on disease risk is independent of the platform used for protein abundance measurement and pQTL identification. Ninety-eight out of 122 associations (76%) were based on IVs from the Olink platform, of which 57 (58%) could be tested using a pQTL from both Olink and SomaScan platforms; 33 of these (58%) were statistically significant in both platforms (5% FDR; **Supplementary** Fig. 1). While overall there was broad agreement between pQTLs from the two platforms, 4 of the 33 statistically significant associations (12%) across both platforms were not directionally consistent (*BCAM* and *PILRA* for AD, and *CD58* and *KLRB1* for MS). We also observed that 9 protein–disease associations reached statistical significance only when the lead *cis* pQTL from the SomaScan platform was used but not when the lead *cis* pQTL from the Olink platform was used (*CTSH* and *LILRB1* for AD, *IDUA* for PD, *CD200*, *INHBC*, *THSD1* and *AHSG* for MS, and *IDUA* and *INHBC* for ALS).

### Sensitivity analysis for Alzheimer’s disease

The GWAS by Bellenguez et al^8^ used proxy cases for AD, where possible cases identified through self-reported family history of dementia were included; these contributed 42% of AD cases, with the balance being physician-diagnosed cases of AD.^8^ As a sensitivity analysis, we used the GWAS by Kunkle et al^41^ to validate that the proteins associated with AD are specific for AD, as it is the latest GWAS for AD that does not include proxy cases.^41^ Out of 54 protein–disease associations, 34 (63%) were associated with AD in the MR analysis using the GWAS by Kunkle et al at 5% FDR (**Supplementary Fig. S2**). All but one (*EPHX2*) of the validated associations were directionally consistent across the two GWAS for AD. The non-validation of some of the proteins could be explained by either the smaller sample size of the Kunkle et al study (i.e., loss of statistical power), or by the misclassification of other causes of dementia as AD in the Bellenguez et al study.

### Co-localisation between plasma protein abundance and neurological diseases

To examine whether confounding by LD can explain the observed associations, we performed co-localisation of the association signals for protein and disease, assuming a single causal variant at each gene locus (**Supplementary Table S4**).^39^ Among the 122 protein-disease pairs, 46 (38%) had strong evidence of co-localisation (posterior probability >0.80), and 27 additional protein-disease pairs (22%) showed moderate evidence of co-localisation (0.60 < posterior probability <0.80). *CR1* co-localised with both AD and MS. We prioritised these 45 proteins, corresponding to 46 protein-disease associations, for further analyses (**Table 2**). Twenty-three out of these 46 protein-disease pairs (50%) represent previously-unreported genetic associations, since they did not reach genome-wide significance in the disease GWAS, nor have they been otherwise previously reported in the GWAS Catalog (**Table 2**).^26^ While the *cis* pQTL for PRSS8 is a previously identified genome-wide significant association for AD, it is located within a different nearby gene (*BCKDK*).

### Association of plasma mRNA abundance with neurodegenerative diseases

We performed an additional two-sample MR analysis to investigate the association of the abundance in plasma of mRNA encoding the proteins with risk of neurodegenerative diseases using the lead *cis* eQTL reported by the *eQTLGen* consortium.^43^ Out of 122 protein-disease associations, a plasma *cis* eQTL for use as an IV was available for 101 (83%) (**Supplementary Table S5**). Of these, 54 (53%) were significantly associated with disease risk at 5% FDR. When we compared the associations using plasma pQTLs and plasma eQTLs (**Supplementary Fig. S3**), we found inconsistencies in the direction of effect in 9 out of 54 cases (17%). The proteins with an inconsistent effect were CD55, TREML2, CEACAM19 and UBASH3B for AD, TPP1 for ALS, and FCRL1, IL2RA, PTPRC and CD58 for MS. Eleven out of 54 associations (20%) also showed evidence of co-localisation across the locus between mRNA abundance and disease risk (**Supplementary Table S6**). Eight proteins showed co-localisation with a neurological disease using both plasma protein abundance and plasma mRNA abundance (*SIRPA* and *CD33* with AD, and *ASF1A*, *VEGFB*, *TYMP*, *PVALB*, *LMAN2* and *CD5* with MS). The remaining three proteins (*ACE* with AD, and *PLEK* and *FCRL3* with MS) showed evidence of co-localisation with plasma mRNA abundance but not with plasma protein abundance. There is strong evidence for distinct causal variants between ACE protein abundance and AD (posterior probability for H3 = 1.00), and moderate evidence for shared causal variants of PLEK and FCRL3 protein abundance with MS (posterior probability for H4 = 0.74 and 0.76, respectively).

### Association of brain mRNA abundance with neurodegenerative diseases

As an additional step to aid interpretation of our findings, we used data on genetic associations with mRNA abundance in four brain regions (i.e., cortex, basal ganglia, hippocampus, and cerebellum) and the spinal cord, as provided by the *MetaBrain* consortium.^53^ For each protein, we selected the lead *cis* eQTL per tissue as an IV, testing a total of 225 mRNA abundance–disease associations (105 in cortex, 78 in cerebellum, 19 in basal ganglia, 14 in hippocampus and 9 in spinal cord), of which 127 (56%) were statistically significant at 5% FDR (**Supplementary Table S7**). When we compared these associations with the associations using the lead plasma protein *cis* pQTL, we found that 96 (76%) were consistent in the direction of effect (**Supplementary Fig. S4**).

Thirty-one out of these 127 associations (24%) showed strong evidence of co-localisation (**Supplementary Table S8**). Two proteins (CR1 with AD in basal ganglia, cortex and hippocampus, and GRN with AD in cerebellum, cortex and hippocampus) were found to have evidence for co-localisation with disease risk from mRNA abundance in three brain regions, while a further 5 proteins (ACE, CD33, SIRPA and PVR with AD in cerebellum and cortex, and LRRC37A2 with AD in cortex and hippocampus) have support for co-localisation with mRNA abundance in two brain regions. Of note is that eight proteins did not show evidence of co-localisation with plasma protein abundance, but they co-localised with mRNA abundance in one or more brain regions (UBASH3B, NSF, LRRC37A2, BIN1 and ACE with AD, and PTPRJ, PLEK, and FCRL3 with MS).

### Association of plasma protein abundance with brain imaging traits

To assess whether the identified proteins might have an impact on brain structure, we examined the association of pQTLs for the 45 proteins with twelve brain-imaging traits (i.e. intracranial brain volume, mean cortical thickness and surface area, 8 subcortical brain volumes, and white matter hyper-intensities). Out of 526 tested associations, 50 (9.5%) were nominally significant at *P* <0.05, but only six (1.1%) were significant at 5% FDR (**Table 3** and **Supplementary Table S9**). All of these latter associations were supported by co-localisation (**Supplementary Table S10**). Plasma APOE abundance was associated with hippocampal volume, amygdala volume, nucleus accumbens volume, and white matter hyper-intensities. Plasma PILRA and PILRB abundance were associated with caudate nucleus volume.

**Table 3.**
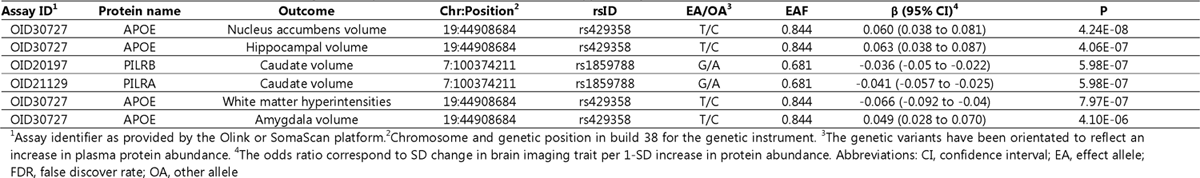
Proteins associated with brain volumes using two-sample Mendelian randomization and supported by co-localisation.

### Assessment of cumulative evidence from protein and mRNA abundance

Our study has combined evidence from MR and co-localisation to identify potentially causal relationships between proteins and neurodegenerative diseases using genetic associations with plasma protein abundance, plasma mRNA abundance, and brain and spinal cord mRNA abundance. On the basis of the overall evidence for association with neurological diseases, we identified three tiers of evidence (**Figure 4**). In Tier 1, we identified 15 proteins (33%) that showed evidence of association and co-localisation when we used a plasma pQTL and an eQTL in at least one brain region. These proteins are TMEM106B, SIRPA, PRSS8, GRN, CR1, CD33, CD2AP and BLNK for AD, HIP1R for PD, TPP1 and TNFSF13 for ALS, and STAT3, PVR, CR1 and ASF1A for MS. In Tier 2, we identified four proteins (8%) that showed evidence of association and co-localisation when we used a plasma pQTL and plasma eQTL (but not studied or detected in any brain region). In Tier 3, we identified 27 proteins (59%) that showed association and co-localisation only when we considered a plasma pQTL but not a plasma or brain eQTL.

**Figure 4.**
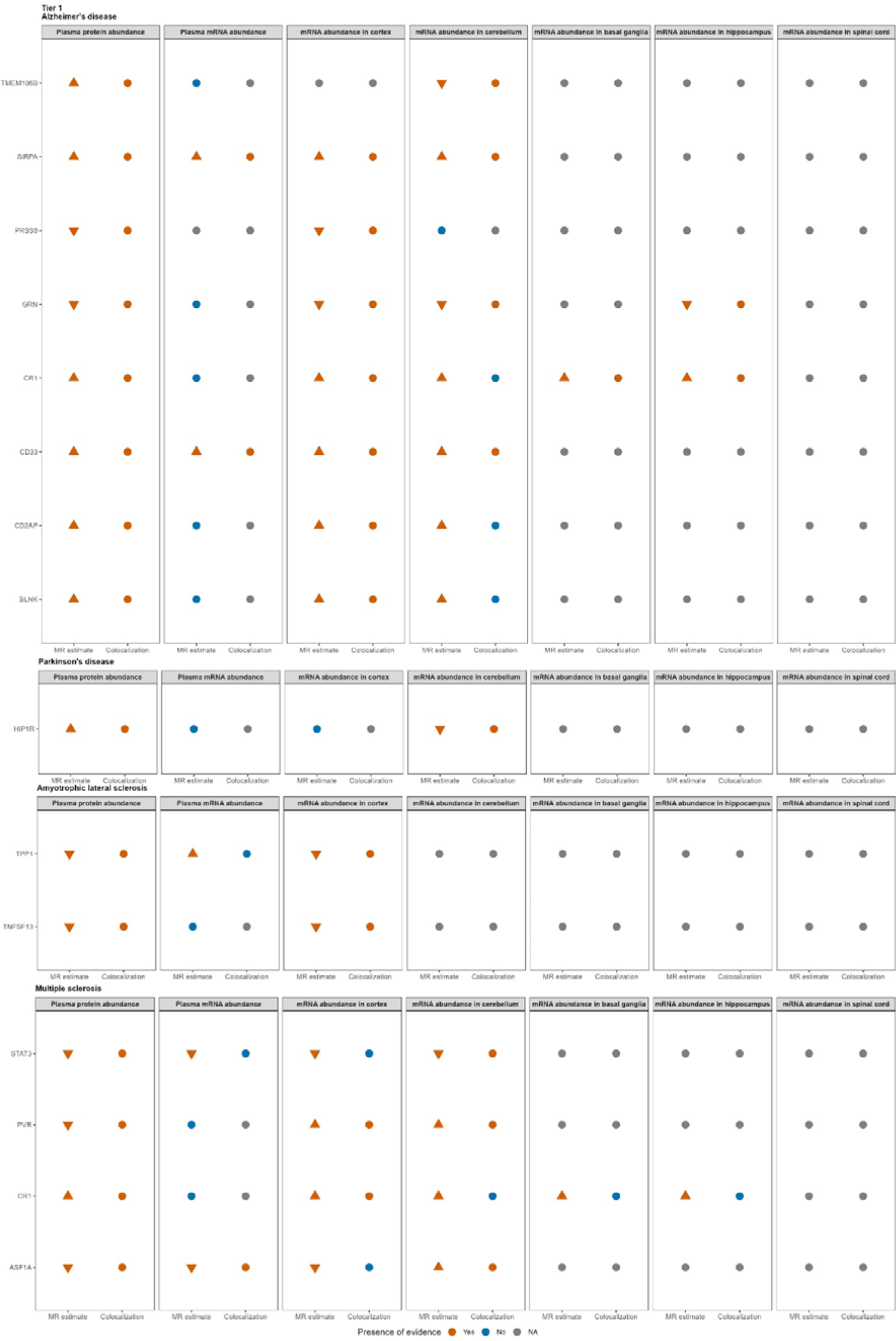
Summary of the evidence using plasma pQTLs and plasma or brain eQTLs as instrumental variables. An eQTL in spinal cord was not available for any of the proteins shown in the figure. When the Mendelian randomisation analysis did not show a statistically significant effect, co-localisation was not performed.

## Discussion

This study has systematically assessed the associations of more than 2,700 proteins with four neurodegenerative diseases using summary statistics from large-scale proteogenomic data and the latest GWAS for disease risk. We identified 46 associations between plasma protein abundance and neurodegenerative diseases with support from MR and evidence of co-localisation. Twenty-three of these associations are known disease loci reported in GWAS, including *APOE*, *MME*, *CD2AP*, *CD33* and *IL34* for AD, and *CD40*, *CD58*, *EVI5*, *IL7R* and *STAT3* for MS, and 23 associations represent previously unreported genetic associations with neurodegenerative diseases.

### Proteins related to AD

β-amyloid accumulation is the main pathogenetic mechanism for AD. APOE is a protein directly involved in the regulation of the β-amyloid aggregation and clearance in the brain.^54^ CD2AP actively participates in the metabolism of β-amyloid, and knockout of *CD2AP* results in endosomal accumulation of β-amyloid in animal models.^55^ MME is also another important enzyme of β-amyloid degradation.^56^

Our findings highlight the role of microglia, which is a cell type equivalent to peripheral macrophages in the brain responsible for the clearance of β-amyloid peptides.^57^ Of note is that one-third of the proteins co-localised with AD relate to microglial function, and our analysis contributes three newly-reported AD loci (*SIRPA*, *SIGLEC9*, *PRSS8*). *CD33* and *IL34* are expressed in microglia and inhibit the microglial uptake of β-amyloid and, therefore, influence the accumulation of amyloid plaque.^58,59^ Anti-CD33 antibodies are used for the treatment of acute myeloid leukaemia and have been previously suggested for drug repurposing for AD.^60^ SIGLEC9 participates in the immune response to several bacterial pathogens by reducing bacterial dissemination into the brain and exerts neuro-protective effects by suppressing inflammatory responses to the brain.^61^ SIRPA regulates microglial phagocytosis, and the transmigration of monocytes across the blood-brain barrier and participates in the pathogenesis of neurodegeneration in preclinical models.^62,63^ *PRSS8* modules Toll-like receptor 4 which is a receptor in the membrane of microglia and contributes to microglial activation and phagocytosis of β-amyloid.^64,65^ The complement system regulates microglial function and neuro-inflammation,^66^ and we identified one known locus (*CR1*) and two newly-identified loci (*C1R* and *C1S*) for AD.

Herpes simplex virus-1 (HSV-1) has been linked with neurodegeneration and cognitive defects in mouse animal models.^67,68^ HSV-1 binds to PILRA, a protein associated with AD, to infect cells. PILRA is a cell surface inhibitory receptor expressed on innate immune cells, including microglia.^69,70^ Also, PILRA was associated with caudate nucleus atrophy, which has been previously observed in other neurodegenerative diseases, including frontotemporal dementia, PD and Huntington disease.^71–73^ This finding could indicate that HSV-1 participates in the pathogenesis of AD by affecting caudate nucleus.

There is an increasing amount of evidence supporting the role in neurodegenerative diseases of lysosomes,^74^ which play an important role in phagocytic cells including microglia.^75^ Our analysis identified one known (*GRN*) and two novel AD-associated loci (*TMEM106B*, *CLN5*) related to lysosomal functions. GRN protein protects against β-amyloid deposition and toxicity in AD mouse models, and its deficiency has been linked with neural circuit development and maintenance, stress response, and innate immunity.^75,76^ TMEM106B has been previously linked with frontotemporal dementia, and there is evidence of its interaction with GRN; both of them are considered critical markers of brain ageing.^77^ Moreover, genetic deficiency of either CLN5 or GRN is responsible for an inherited lysosomal disease.^78,79^ Loss of CLN5 leads to deficits in neurodevelopment in mice models.^78^

Our analysis identified three additional newly-identified proteins (PLOD2, ZBTB18, BLNK) potentially participating in the pathogenesis of AD through other pathways. PLOD2 is overexpressed in fibroblasts, strengthening the current evidence for a potential role for fibroblasts in the pathogenesis of AD through remodelling of the extracellular matrix alongside amyloid plaques.^80^ ZBTB18 is an essential transcription factor for embryonic cerebral cortex development;^81^ it has been identified as a contributing factor to the 1q43q44 microdeletion syndrome, which is characterised by variable intellectual disability and brain malformations.^82^ BLNK is involved in B cell receptor signalling; although the role of B cells in AD is not well understood, targeting B cells has been suggested to be beneficial for AD patients by delaying disease progression.^83^

### Proteins related to PD and ALS

Although the GWAS for PD and ALS have relatively small sample sizes, our study was nevertheless able to identify one novel locus for PD (*CTF1*) and two novel loci for ALS (*TPP1*, *TNFSF13*). CTF1 is a neurotrophic factor in the interleukin-6 cytokine family. Pro-inflammatory cytokines, including interleukin-6, have been previously associated with PD.^84^ It has also been shown in a mouse model that *CTF1* transfection and expression is neuroprotective and slows progression of spinal muscular atrophy.^85^ One of the aetiologies of ALS is mis-localization of TDP-43 to mitochondria causing neurotoxicity.^86^ TPP1 is a lysosomal enzyme and loss-of-function mutations in the gene are causally linked to a familial lysosomal disorder, in which *TPP1* loss affects regulation of axonal mitochondrial transport.^87^ Also, loss of TPP1 activity results in progressive neurological phenotypes including ataxia and increased motor deficiency.^88^ *TNFSF13* is expressed in astrocytes and regulates neuro-inflammatory responses.^89^ Reactive astrocytes have neurotoxic properties and are involved in the pathogenesis of ALS.^90^

### Proteins related to MS

Experimental autoimmune encephalomyelitis (EAE) is an animal model for MS, and 9 of the MS-associated proteins, 6 known (CD5, CD40, IL7R, STAT3, TNFRSF1A, TYMP) and 3 newly-associated with MS (PARP1, PVALB, VEGFB), are involved in the pathogenesis of EAE. This observation strengthens the validity of our findings.^91–99^

The innate immune system participates in pathogen removal and regulates the response of the adaptive immune system, ^100^ including the response to Epstein-Barr virus (EBV) infection which is a pathogen associated with MS.^101^ *PVR*, a known loci for MS, encodes the polio virus receptor, which is involved in the immune response to EBV. Increased expression of *PVR* downregulates the expression of miRNAs produced by EBV,^102^ which potentially explains the apparent protective effect of higher plasma levels of PVR in our analysis. WARS1, a protein newly-associated with MS, is an aminoacyl-tRNA synthetase with a role as an innate immune activator in the extracellular space, acting as a primary defense system against infections and especially antiviral immunity.^103,104^ Moreover, a newly-identified MS protein, PARP1, is involved in the NF-κB signaling pathway,^105^ which is activated as a response to infectious antigens including EBV.^106^ and is an important pathway for the activation of macrophages and other innate immune cells.^107^ The newly-identified association of CR1 with MS risk indicates a role of complement, which is an important innate immune defence against infection, as has been recently suggested.^108^ SLAMF1 participates in the Toll-like receptor 4 signalling which activates macrophages against bacterial pathogens.^109^ This observation potentially provides support to the hygiene hypothesis for the development of MS.^110^

The adaptive immune system consists of B cells and T cells, which are activated by innate immune cells, IL7R has a role in T and B cell differentiation, and its plasma levels are associated with elevated risk of MS,^111^ but experimental IL7R inhibitors have not been successful in treating MS.^112^ CD5 and CD58 are also involved in B and T cell differentiation and whose activation has a role in autoimmunity.^113,114^ CD40 and its ligand form a complex that has a central role in the regulation of both humoral and cell-mediated immunity. Blockade of CD40L is effective in ameliorating experimental autoimmune conditions, and it has also been suggested as a potential therapeutic strategy for MS.^115^

Demyelinating lesions in white and grey matter are the histopathological landmark of MS, which are infiltrated by cells of the innate and adaptive immune system,^100^ whereas oligodendrocytes are responsible for the myelination process.^116^ PARP1, a newly-associated protein for MS, is a driver for oligodendroglial development and myelination,^117^ and PARP1 inhibitors have been suggested as a potential therapy for MS,^118^ in line with our finding that elevated plasma PARP1 is associated with increased MS risk. STAT3, a known protein for MS, is important for myelin repair, and pharmacological blockade of STAT3 activation with JAK2 inhibitors inhibits survival and differentiation of oligodendrocyte precursor cells.^119^ Another known protein for MS, TNFRSF1A, is involved in the TNF receptor-associated periodic syndrome, which is characterised by inflammatory demyelination. There is evidence that anti-TNFα therapies can result in new episodes of inflammation in MS patients.^120^

The blood-brain barrier (BBB) protects the central nervous system parenchyma from harmful circulating molecules and pathogens,^121^ and altered BBB function is believed to be an important early stage in MS pathology. Several identified proteins have potential roles in the BBB, including TYMP, a key astrocyte-derived permeability factor promoting BBB breakdown,^92^ CD40, which influences the permeability of the BBB,^122^ and VEGFB, a newly-identified MS-associated protein which is a member of the vascular growth factor family, again involved in the permeability of the BBB.^123^

The node of Ranvier on white matter demyelinated axons is profoundly altered or disrupted in patients with MS,^124,125^ and two newly-identified proteins (NCS1, CDH15) are involved in its function. NCS1 is involved in the regulation of intracellular calcium signalling and is identified in the nodes of Ranvier. NCS1 also participates in the pathogenesis of chemotherapy-induced peripheral neuropathy.^126^ A member of the cadherin protein family, CDH15, participates in the function of the node of Ranvier and has previously been associated with chronic inflammatory demyelinating polyneuropathy, a demyelinating disease of the peripheral nervous system.^127^ Lesions in the dorsal root ganglion are identified in EAE,^128,129^ and *FLRT3*, a newly-identified protein related to MS, is overexpressed in the dorsal root ganglion and has been associated with neuropathic pain in animal models.^130^

Four further newly-identified MS-associated loci (*PVALB*, *TST*, *ASF1A*) potentially indicate additional molecular pathways contributing to MS. PVALB is specifically expressed by GABAergic interneurons and has been suggested as a potential MS-specific marker of grey matter neuro-degeneration.^131^ TST is an enzyme involved in mitochondrial sulphur and selenium metabolism,^132^ and it has been shown that exposure to oxidative stress due to mitochondrial dysfunction contributes to the chronic demyelination.^133^ ASF1A is a histone chaperone which has been implicated in neuro-inflammation and neuro-degeneration processes through activation of microglia.^134,135^

### Plasma and tissue-specific proteomic effects

The identification of proteins with roles in many of the biological processes relevant to neurodegenerative diseases supports the idea that targeting such proteins might form the basis of future drug development. However, it seems likely that abundance of these proteins in blood plasma is not directly relevant to disease pathology, and that therapies will need to be targeted to the relevant tissue or cell-type. Even though a drug may modify levels of the identified proteins in plasma, we cannot assume that it would cross the blood-brain barrier for brain-targeting drugs.^136^ Nevertheless, although our results are primarily based on proteins measured in plasma, it is plausible that the same genetic factors have similar effects on protein levels in more relevant tissues, and that our results reflect similarities in processes such as macrophage activity, lysosomal activity, and β-amyloid metabolism in blood and brain. For example, in the pathogenesis of MS, the activation of innate and adaptive immune system occurs first in the periphery and is then transferred in the central nervous system.^100^ In particular, *cis* pQTLs, particularly those directly impacting protein coding sequences, will frequently have similar effects across diverse tissues.^137^

### Limitations

There are several limitations to the present study. First, our analyses are underpowered for PD, because full summary statistics including 23andMe are not publicly available, greatly reducing the sample size in the available data. Second, GWAS for the neurological diseases primarily assess risk for disease and not disease progression. Therefore, the identified proteins can be considered as potential biomarkers for prediction or diagnosis of the diseases or drug targets for disease prevention but not necessarily for disease progression.^138^ Third, effect estimates based on MR assume a lifelong exposure to altered protein levels. For this reason, MR estimates frequently differ from observational associations and do not necessarily reflect the impact of a change in protein levels as a result of a therapeutic intervention.

## Conclusions

To summarise, we presented a comprehensive analysis of associations of the plasma proteome with neurodegenerative diseases by considering proteins measured through either Olink or SomaScan platforms. We identified multiple proteins with potential causal role in neurodegenerative diseases. The newly-identified proteins for AD are involved in the immune response to bacterial pathogens, complement system, transmigration of monocytes across the blood-brain barrier, Toll-like receptor 4 signalling, lysosomal function and fibroblasts. The newly-identified proteins for MS are involved in the innate immune system, complement, microglia, oligodendrocytes, permeability of the BBB, GABAergic interneurons and the function of node of Ranvier and dorsal root ganglion. Our analysis covered only a modest proportion of the human proteome, and was limited to proteins measured in blood plasma; therefore, further expansion of the multiplexed antibody-based and aptamer-based assays, and conducting large-scale assays in more directly-relevant tissues, will offer additional insights into the role of protein abundance on the development of neurodegenerative diseases. Moreover, better characterisation of the protein isoforms targeted by these complementary proteomics platforms will offer additional insights into the biological interpretation of the findings.

## Abbreviations

AD: Alzheimer’s disease

ALS: amyotrophic lateral sclerosis

BBB: blood-brain barrier

eQTL: expression quantitative trait locus

FDR: false discovery rate

IV: instrumental variable

LD: linkage disequilibrium

GWAS: genome-wide association study

MHC: major histocompatibility complex

MR: Mendelian randomization

MS: multiple sclerosis

PD: Parkinson’s disease

pQTL: protein quantitative trait locus

WMH: white matter hyper-intensities

## Data availability

The *cis* pQTLs that were used as IVs are publicly available in the relevant publications.^25,51^ Summary statistics for the GWAS on AD and ALS are available through GWAS Catalog (https://www.ebi.ac.uk/gwas/). Summary statistics for the GWAS on PD and MS are available through OpenGWAS project (https://gwas.mrcieu.ac.uk/). Summary statistics for the GWAS on brain volume traits are available upon request from the ENIGMA consortium (https://enigma.ini.usc.edu/). Summary statistics for the GWAS on white matter hyper-intensities are publicly available through CHARGE consortium. Summary statistics for the *cis* region of gene expression in plasma and brain regions are publicly available through the eQTLGen (https://www.eqtlgen.org/) and the MetaBrain (https://www.metabrain.nl/) consortia, respectively.

## Acknowledgements

A preliminary version of this research article was presented as a poster at the 2023 Annual Meeting of the American Society of Human Genetics.

## Supplementary material

Supplementary material is available at *Brain* online.

