## Supplementary material 1 for "A Mendelian randomization study identifies proteins involved in neurodegenerative diseases"

### Supplementary text

#### Estimation of F-statistic

F-statistic was calculated for each SNP using the following formula:

$$F = \frac{R^2 \times (N - 2)}{1 - R^2}$$

where $R^2$ is the proportion of variance explained by each SNP, and $N$ is the sample size of the GWA study. $R^2$ was calculated for each SNP using the following formula:

$$R^2 = \frac{2 \times EAF \times (1 - EAF) \times beta^2}{[2 \times EAF \times (1 - EAF) \times beta^2] + [2 \times EAF \times (1 - EAF) \times N \times SE(beta)^2]}$$

where $EAF$ is the effect allele frequency, $beta$ is the estimated genetic effect on the protein abundance, $N$ is the sample size of the GWA study, and $SE(beta)$ is the standard error of the estimated genetic effect on the protein abundance.

#### Estimation of effect estimate from Z-score

In the summary statistics of brain phenotypes from the ENIGMA consortium, the results are as Z-scores and P-values. Effect estimates and standard errors were derived using a formula from the literature:[136]

$$beta = \frac{z}{\sqrt{2p(1 - p)(n + z^2)}}$$

$$se = \frac{1}{\sqrt{2p(1 - p)(n + z^2)}}$$

where $p$ is the minor allele frequency of the genetic variant and $n$ is the sample size.

### Supplementary Figures

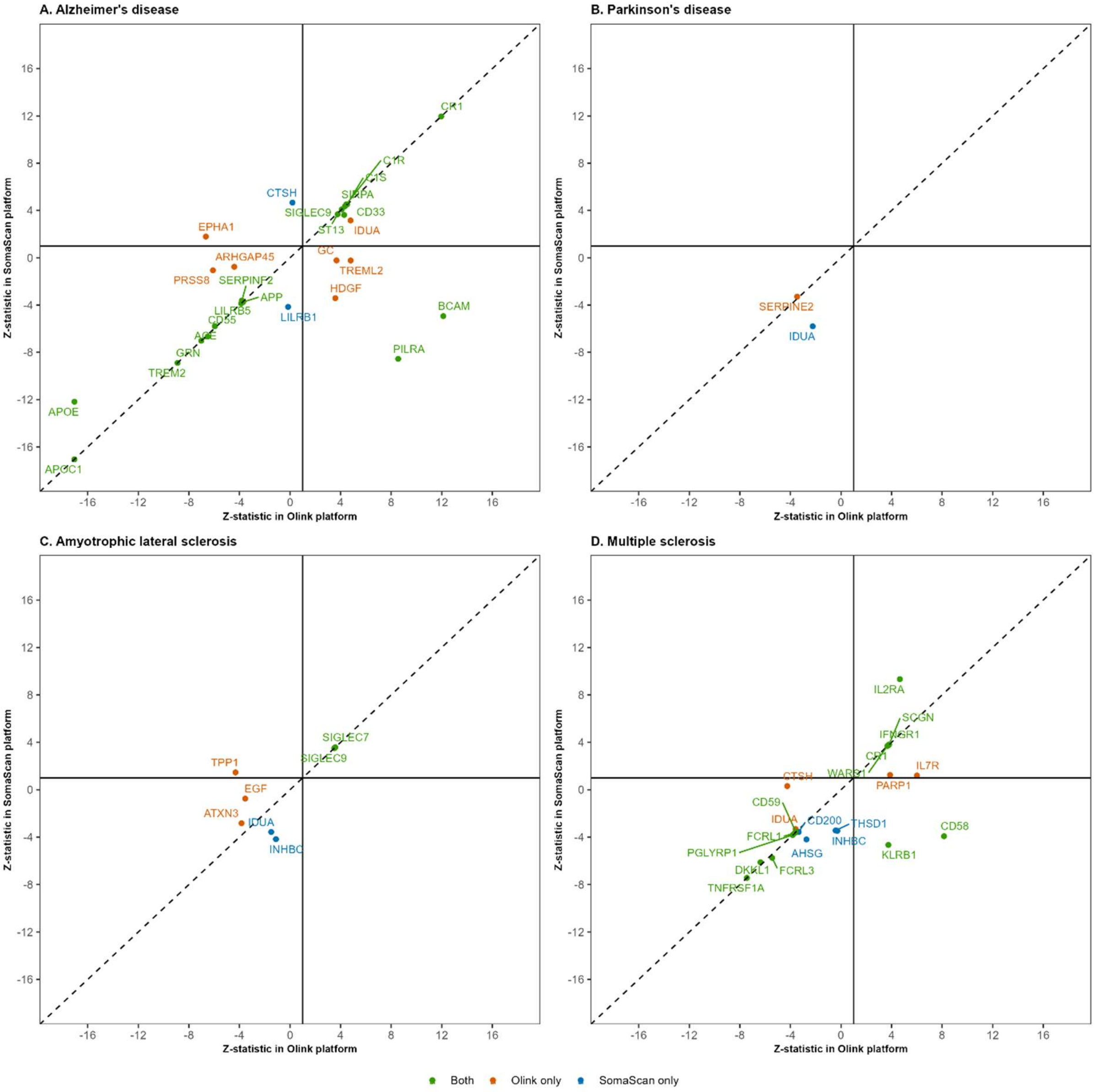


**Supplementary Fig. S1. Comparison of the MR effect estimate of plasma protein abundance on neurological diseases between the Olink and SomaScan platform.** For proteins that showed a statistically significant effect on neurological diseases using the lead *cis* pQTL from the Olink platform, we repeated the MR analysis using the lead *cis* pQTL from the SomaScan platform, when available. The scatter plots show the correlation of Z statistic for the effect estimates between the Olink and the SomaScan platforms. Proteins that were significant using either a pQTL from Olink or SomaScan are shown in green, proteins that were significant only using a pQTL from Olink are shown in orange, and proteins that were significant only using a pQTL from SomaScan are shown in blue.

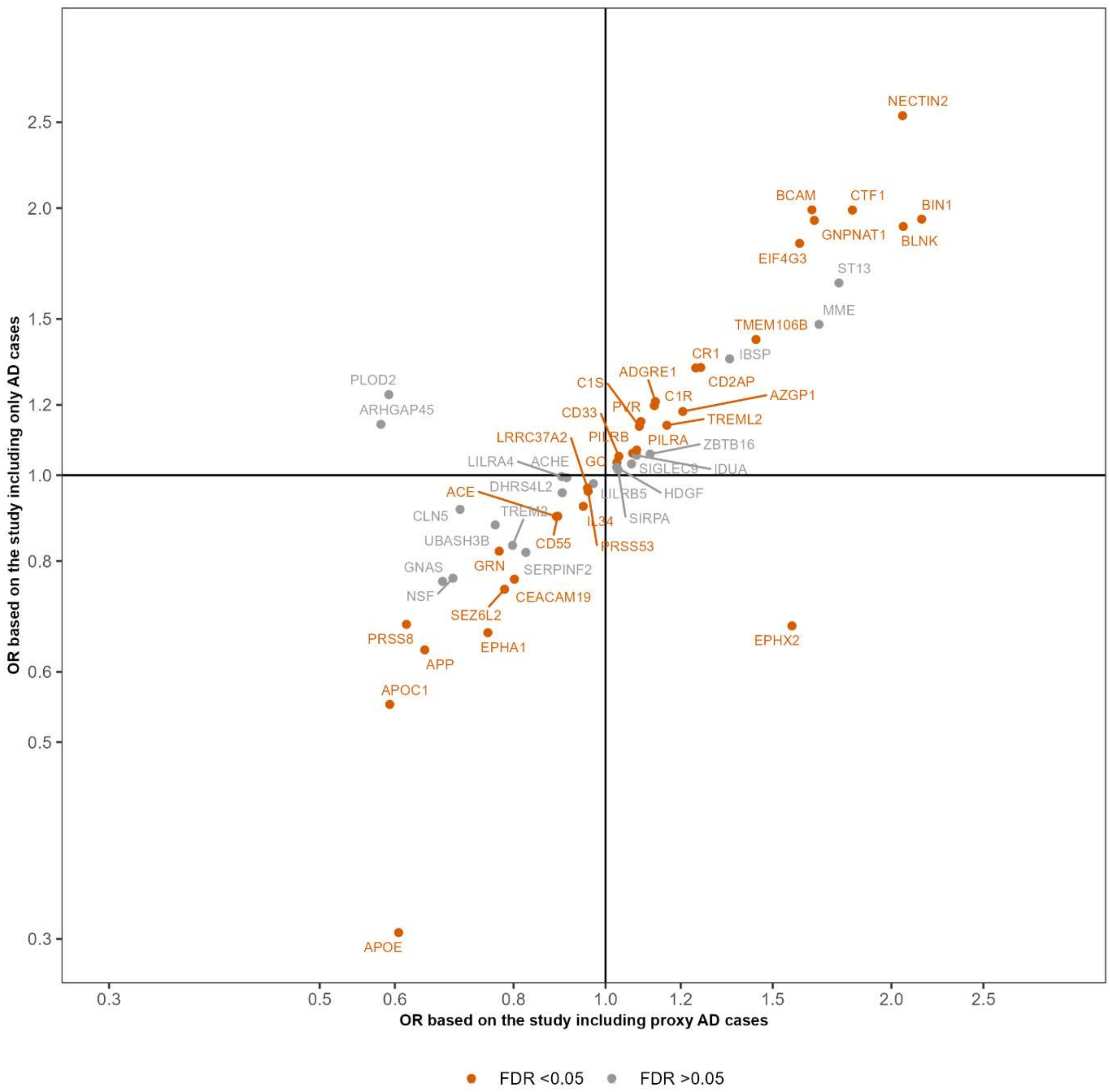


**Supplementary Fig. S2. Comparison of the MR effect of plasma protein abundance on Alzheimer's disease using two different genome-wide association studies for Alzheimer's disease.** All the proteins shown in the plot are statistically significant for an association with Alzheimer's disease when we use the GWAS including physician-diagnosed AD cases and proxy AD cases to derive the gene – outcome effect for the Wald ratio. The color-coded legend is based on statistical significant at 5% false discovery rate when we use the GWAS including only physician-diagnosed cases to derive the gene – outcome effect.

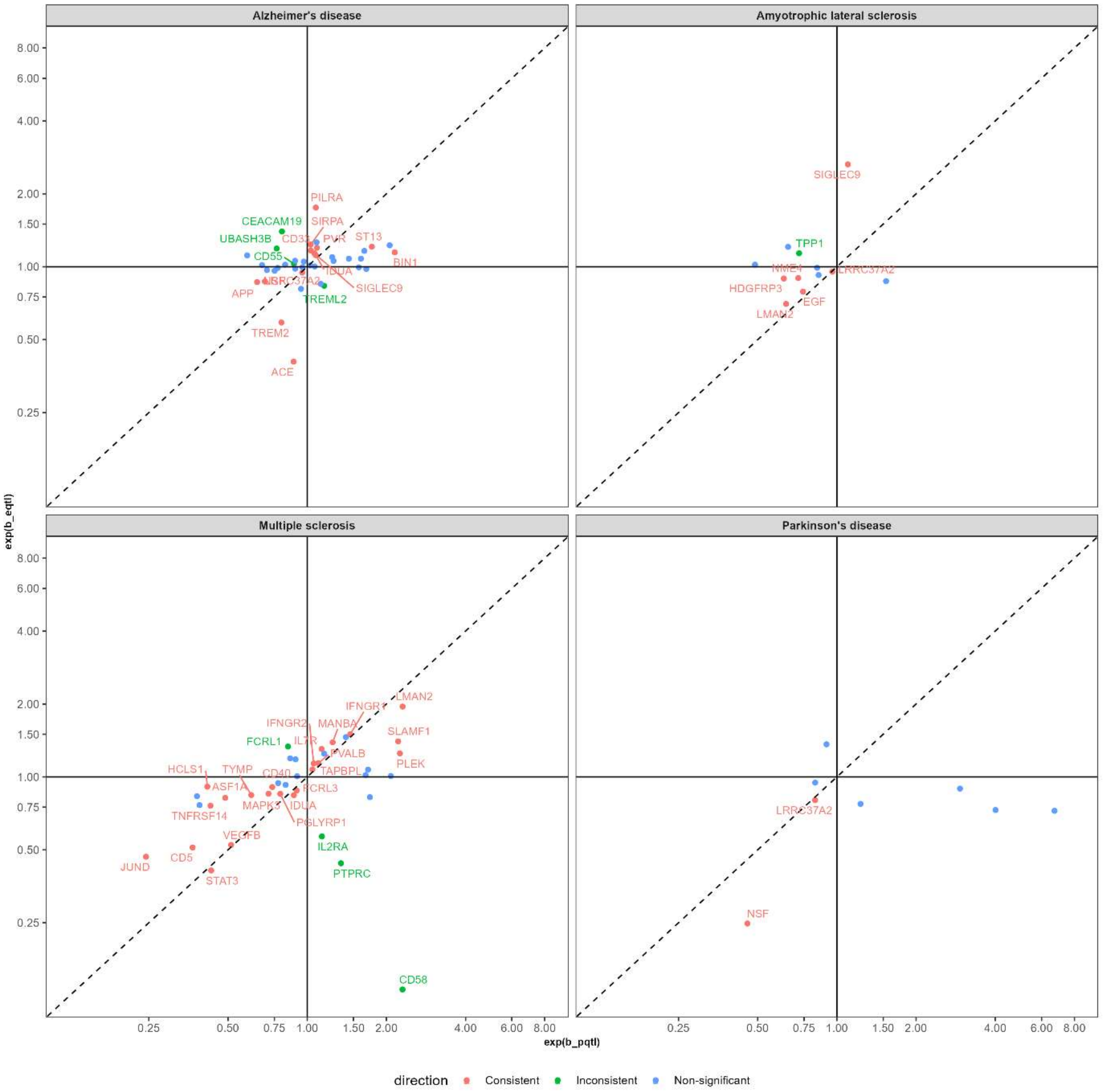


**Supplementary Fig. S3. Comparison of MR effect estimate of plasma protein abundance and plasma transcript abundance on neurological diseases.** The color-coded legend is based on the consistency of the effect estimate using a plasma pQTL and plasma eQTL.

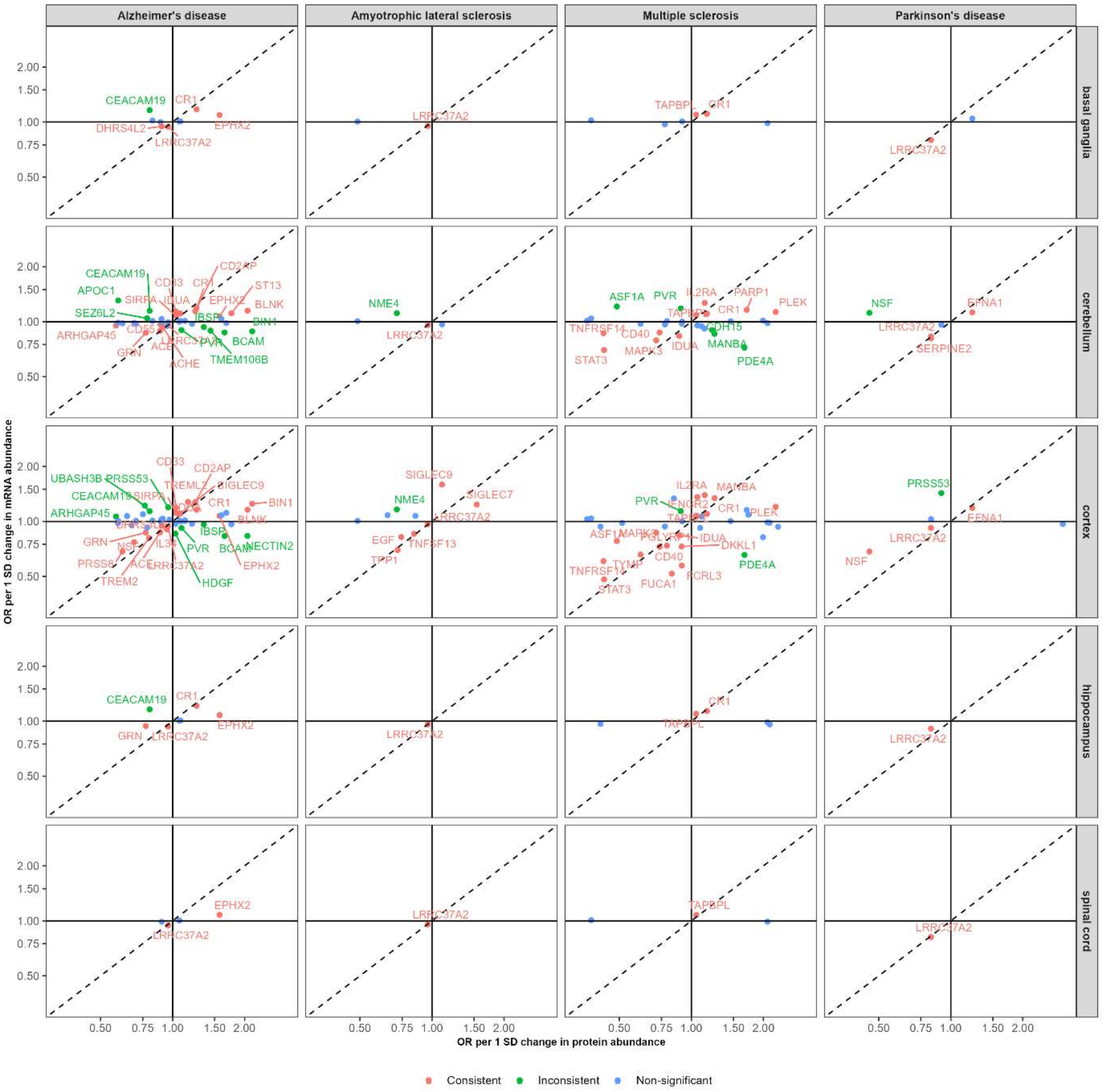


**Supplementary Fig. S4. Comparison of the MR effect estimate of plasma protein abundance and transcript abundance in brain on neurological diseases.** The color-coded legend is based on the consistency of the effect estimate using a plasma pQTL and brain eQTL.
